# Harmonized US National Health and Nutrition Examination Survey 1988-2018 for high throughput exposome-health discovery

**DOI:** 10.1101/2023.02.06.23284573

**Authors:** Vy Kim Nguyen, Lauren Y. M. Middleton, Lei Huang, Neil Zhao, Eliseu Verly, Jacob Kvasnicka, Luke Sagers, Chirag J. Patel, Justin Colacino, Olivier Jolliet

**Author notes:** Corresponding author: Vy Kim Nguyen, PhD ( or).

## Abstract

The National Health and Nutrition Examination Survey (NHANES) provides data on the health and environmental exposure of the non-institutionalized US population. Such data have considerable potential to understand how the environment and behaviors impact human health. These data are also currently leveraged to answer public health questions such as prevalence of disease. However, these data need to first be processed before new insights can be derived through large-scale analyses. NHANES data are stored across hundreds of files with multiple inconsistencies. Correcting such inconsistencies takes systematic cross examination and considerable efforts but is required for accurately and reproducibly characterizing the associations between the exposome and diseases. Thus, we developed a set of curated and unified datasets and accompanied code by merging 614 separate files and harmonizing unrestricted data across NHANES III (1988-1994) and Continuous (1999-2018), totaling 134,310 participants and 4,740 variables. The variables convey 1) demographic information, 2) dietary consumption, 3) physical examination results, 4) occupation, 5) questionnaire items (e.g., physical activity, general health status, medical conditions), 6) medications, 7) mortality status linked from the National Death Index, 8) survey weights, 9) environmental exposure biomarker measurements, and 10) chemical comments that indicate which measurements are below or above the lower limit of detection. We also provide a data dictionary listing the variables and their descriptions to help researchers browse the data. We also provide R markdown files to show example codes on calculating summary statistics and running regression models to help accelerate high-throughput analysis and secular trends of the exposome.

|  |  |
| --- | --- |
| Measurement types | demographics, diet, health, physiological measurements, and blood and urine exposure biomarkers |
| Technology types | interviews, examination, laboratory tests |
| Sample characteristic – organization | homo sapiens |
| Sample characteristic – location | United States |

**Background & Summary:** The Centers for Disease Control and Prevention (CDC) designed the National Health and Nutrition Examination Survey (NHANES) to monitor the health and nutritional status of adults and children in the US^1^. Beginning in 1960s, the NHANES program has included a series of surveys to ascertain health, nutritional, and environmental measurements in nationally representative samples. NHANES III was conducted in 1988-1994, consisting of a total of 39,695 persons aged 2 months and older, and was separated into two phases, Phase 1 (1988-1991) and Phase 2 (1991-1994). Continuous NHANES is a series of surveys conducted every two years since 1999, consisting of approximately 10,000 participants for every two-year study period to culminate to a total of 134,310 participants^2^. The data were ascertained through home interviews and health examinations. The interviews included self-reported questionnaires on demographics, socioeconomic status, health history, dietary intake, and occupation^2^. The health examination was conducted in mobile exam centers (MEC) staffed by a highly trained medical team^2^. The examination consists of clinical and physiological measurements of health metrics. In addition, the physical examination includes laboratory tests to quantify biomarkers of environmental chemical exposures in serum or urine. Overall, NHANES houses a wealth of data to be used to understand the health of the nation, but the potential to study secular changes in associations, such as in cancer mortality, over a wide range of time have not been evaluated.

---

The wealth of data in NHANES can be used to study the “exposome,” which is the totality of all environmental exposures encountered in a human lifespan and estimate how these exposures impact health^3,4^. Conducting exposome analyses on extensive datasets like NHANES will be integral for identifying expected and novel risk factors of disease. But such efforts have been hampered due to inconsistencies in NHANES. Some common inconsistencies in NHANES include changes in variable nomenclature, changes in units of measurements, and changes in labels for the same category. First, an example of changes in variable nomenclature is when the same variable changes its name part way through the study. This may lead to substantial amount of data being excluded from analysis if this change is not taken into account. Moreover, if two completely different variables have the same name, then this mistake would result in erroneous analysis and interpretations, unless it is corrected. Second, changes in the units of measurement can differ by several orders of magnitude. Not accounting for this conversion could lead to mistakenly interpreting substantial changes in the variable over time when there were none. Finally, for categorical variables, changes in labels for the same category could lead to misclassification. Overall, such inconsistencies in NHANES can result in errors when analyzing the data and interpreting the findings. Thus, our goal is to systematically correct for these inconsistencies to create a set of harmonized and unified datasets to decrease the amount of time that researchers spent cleaning and consequentially to help increase time and effort for analysis to accurately arrive at public health and medical insights earlier. Furthermore, creating a single repository of curated datasets will facilitate interdisciplinary approaches as well as exposome approaches to study the interaction and combined effects of the major determinants (e.g., environmental exposures, dietary habits, physical activities) of health.

We expanded upon the work of Patel et al., 2016 on the database of human exposomes and phenomes from the US NHANES 1999-2006^5^ to create a harmonized and unified version that includes 1988-2018. This resource can enable the community to survey temporal or secular based trends in associations between the exposome and mortality, which is a critical question in public health. NHANES can complement the National Cancer Institute Surveillance, Epidemiology, and End Results (SEER) Program^6^ by providing opportunities to associate the exposome with cancer outcomes (e.g., cancer-specific mortality). While the SEER program is a great source for tracking trends in cancer incidence and outcomes in the US^6^, it has been limited in the availability of exposome variables to study the impact of the environment on cancer. Furthermore, we integrated data from NHANES III with those of NHANES Continuous to widen the study period to increase the number of death cases that are ascertained. This wider study period will enable researchers to study how the associations between the environment and cancer outcomes change over time. Relative to Patel et al., 2016, our expansion tripled the population size (N = 134,310 vs. N = 41,474), quadrupled the number of variables (p = 4,740 vs. p = 1,191), increase the number of cases due to all-cause mortality by a factor of 12 (d = 17,872 vs. d = 1,357), tripled the number of death causes (c = 10 vs. c = 3), and nearly tripled the number of survey cycles (s = 11 vs. s = 4). These improvements resulted in greater statistical power to study exposome-mortality risk.

The NHANES data have been commonly used in developing health policy. For example, these data were instrumental in removing lead from gasoline, and since such legislation, there has been a substantial decrease in blood lead levels in the US population^2^. These data have also been used to establish national estimates of the prevalence for elevated cholesterol, elevated blood pressure, and Hepatitis C in the US^2^. These data are also powerful to detect trends in exposure across many classes of chemicals. For example, several studies have used these integrated data to screen across 100-150 environmental toxicants to identify toxicants with elevated exposure in sensitive populations^7–10^. To link to health endpoints, many have used these data to test hypotheses on associations between environmental risk factors and disease, such as heart disease^11,12^ and all-cause mortality^13–15^. Patel et al, 2016 used NHANES to conduct the first “environment-wide association studies” (EWAS) by screening across over 250 chemical biomarkers with respect to diabetes and have made these data broadly available^16^. Thus, we hope that our set of curated NHANES datasets will facilitate large-scale analysis like EWAS^17,18^ and machine learning approaches to better understand the combined influence of the exposures on disease.

In this data descriptor, we provide (1) a dictionary of available variables in our unified NHANES data, (2) tabulated documentation to provide transparency of our curation process, and (3) the curated and unified NHANES datasets as either .csv or .RData files.

## Methods

The NHANES datasets are publicly accessible through www.cdc.gov/nchs/nhanes/index.htm and ftp.cdc.gov/pub/Health_Statistics/NCHS/datalinkage/linked_mortality/. All NHANES participants consented for their information to be used for research. This study was reviewed by the University of Michigan Review Board and deemed to be exempted (HUM00116291). We developed the curation pipeline using R version 4.0. In this section, we describe the 1) procedure of how we downloaded and compiled the datasets, 2) brief description of each type of dataset, 3) the curation procedure for each type of dataset, 4) the creation of the dictionary to peruse through the data, and 5) starter code to conduct mortality-related exposome analysis. Figure 1 shows our workflow.

**Figure 1.**
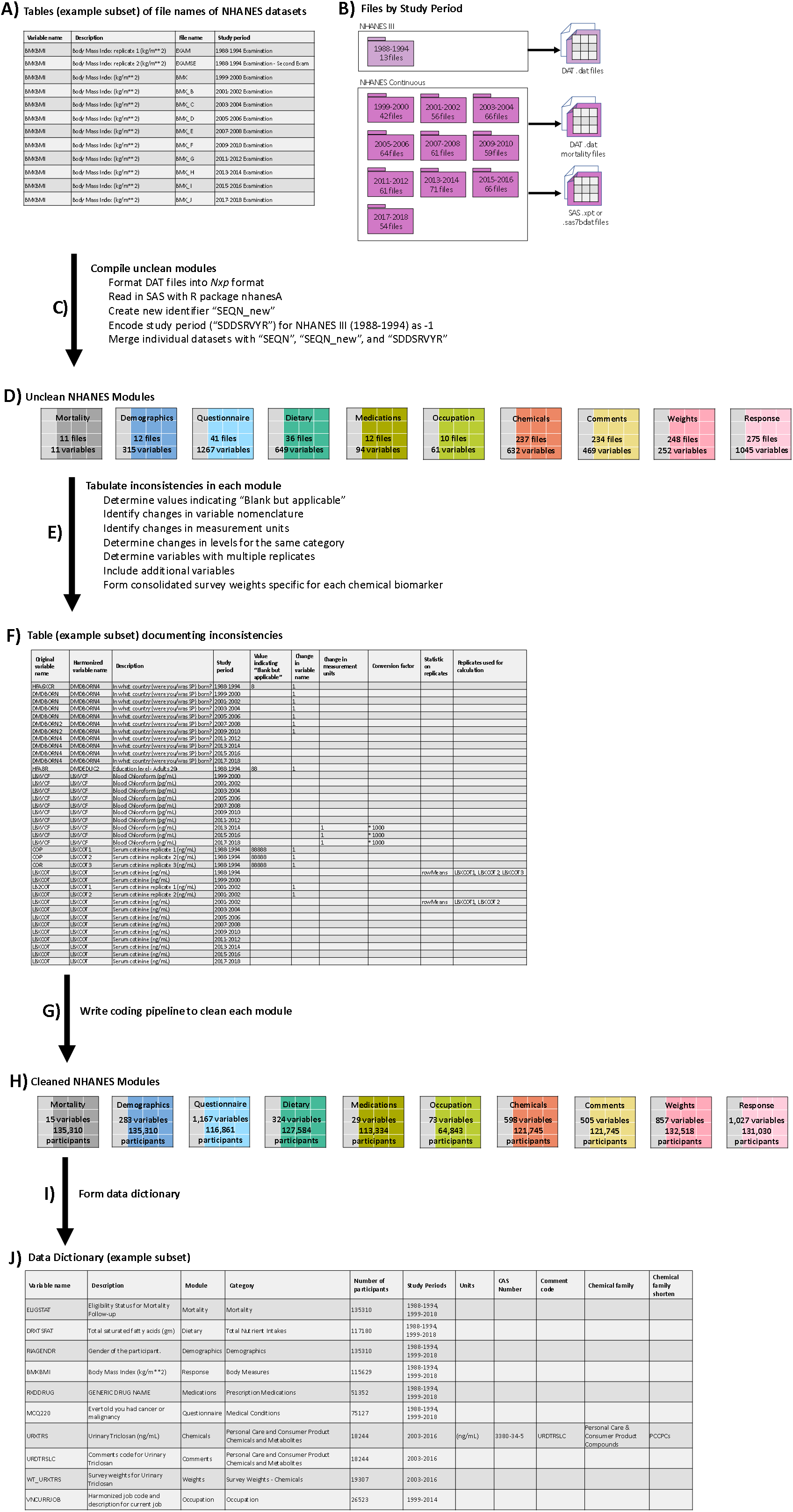
Overview of the created NHANES dataset and workflow schematic for creating and cleaning these datasets. A) List of the variable names and the file name of the NHANES datasets containing the corresponding variables. B) Number of files and types of files (SAS or DAT files), stratified by study period. C) Process to download and compile individual NHANES datasets into the form of Nxp by using the tables of file names. D) Resulting 10 uncleaned modules corresponding to different types of datasets. We listed the number of files used to create each module and the number of variables based on counts of unharmonized variable names. E) Process to detect and tabulate of inconsistencies in each module. F) Table to document the different types of inconsistencies. G) Process to curate each module by using the cleaning documentation. H) List of number of variables and participants for each cleaned module. I) Process to create data dictionary. J) Data dictionary that contains the variable and human readable descriptions along with other meta-data, such as module name, variable category, number of participants, and study periods in which the variable is available. For the chemical variables, this table include additional meta-data such as units of measurement, CAS Number, comment codename, chemical family, and abbreviated name of chemical family.

### 1. Procedure to download files

The CDC provided the NHANES data in many individual datasets, which can be intimidating and daunting to browse through several documentations to determine which variables are in which datasets. Thus, we developed a table for each type of data (e.g. mortality, demographics, etc.) listing the variable names and the corresponding file names of the NHANES datasets housing the variables (example in Figure 1A, Tables S1, S3, S5, S7, S9, S10, S12, S13, S14). We then used these tables of filenames to download a total of 614 files, delineated by study period: 1988-1994 (13 files), 1999-2000 (42), 2001-2002 (56), 2003-2004 (66), 2005-2006 (64), 2007-2008 (61), 2009-2010 (59), 2011-2012 (61), 2013-2014 (71), 2015-2016 (66), and2017-2018 (54) (Figure 1B). There is one file, “Prescription Medications - Drug Information,” that spans across NHANES III (1988-1994) and NHANES Continuous (1999-2018). The CDC ascertained causes and time to death information by linking eligible NHANES participants to the National Death Index. Thus, we downloaded 11 mortality files, which are encoded as DAT “.dat” format. The 13 files from NHANES III are also encoded as DAT, while files from NHANES Continuous are encoded as SAS “.xpt” or “.sas7bdat” format.

We used the following procedure to ensure that the DAT and SAS files are read into R as consistent tables (Figure 1C). We first manually downloaded the DAT files from NHANES III (https://www.n.cdc.gov/nchs/nhanes/nhanes3/datafiles.aspx#core) and from the mortality repository (https://ftp.cdc.gov/pub/Health_Statistics/NCHS/datalinkage/linked_mortality/). DAT files are structured as a series of numbers concatenated into one number per each row (each participant). We reformatted this file as an Nxp table, where N represents the participants and p represents the variables. This requires that we know the start and end position of each variable in the DAT files to concatenate the numbers into a variable. The start and end positions of each NHANES variable can be assessed using the SAS code from https://www.n.cdc.gov/nchs/nhanes/nhanes3/datafiles.aspx#core, and for the mortality variables, the positions can be found in the R code from https://ftp.cdc.gov/pub/Health_Statistics/NCHS/datalinkage/linked_mortality/. We used the readr^19^ package and specified the width of each variable to read the DAT files and transform it into the table form of Nxp. For the SAS files, we directly downloaded the file using the R package nhanesA^20^. When the files are read into R, the format of the SAS files are already in the table form of Nxp. For the NHANES Continuous datasets, the study period (SDDSRVYR) is encoded as 1 for 1999-2000, 2 for 2001-2002, and onward until 10 for 2017-2018. Since NHANES III datasets do not have this variable, we encoded this study period as -1.

NHANES is a cross-sectional and there is no overlap in participants across the study periods. Each participant in NHANES has an identifier, “SEQN”, but this identifier is not any more unique when merging NHANES III and NHANES Continuous, with different participants having the same SEQN (one in NHANES III and another in NHANES Continuous). Thus, we created a new identifier “SEQN_new” to differentiate participants between NHANES III and NHANES Continuous participants, ensuring that each NHANES participant has a unique identifier. To create “SEQN_new”, we concatenated the SEQN number with “III” for NHANES III participants and with “C” for NHANES Continuous participants. Among the different types of datasets, the common variables are “SEQN”, “SEQN_new”, and “SDDSRVYR”. We merged the individual NHANES datasets using “SEQN”, “SEQN_new”, and “SDDSRVYR” (Figure 1C). We involved “SDDSRVYR” in the merging process, because if it was not included, then there would be duplicates of this variable. Multiple duplicates would excessively increase the size of an already large dataset and consume excessive storage. Moreover, the labels of this variable would also become excessive (e.g. “SDDSRVYR.x”, “SDDSRVYR.y”, “SDDSRVYR.x.x”, “SDDSRVYR.y.y”). In total, the 614 data files contain information on 134,310 distinct individuals and 4,685 un-harmonized, original variables.

### 2. Modules and Types of Datasets

We have created modules (or categories of datasets) based on how the CDC organized their data (Figure 1D). We delineate the modules as the following: 1) Mortality, 2) Demographics, 3) Questionnaire, 4) Dietary, 5) Medications, 6) Occupation, 7) Chemicals, 8) Comments, 9) Weights, and 10) Response. The Mortality module contains information on death status as well as on causes of death, followed up through December 31, 2019. The Dietary module houses nutrient information estimated from the foods and beverages consumed by the participants during the 24-hour period before the interview. The Demographic module contains information such as age, sex, race/ethnicity, socioeconomic status, etc. The Response module contains body measurements (e.g., BMI, weight, height, skinfold) and biomarker measurements (e.g., cholesterol, blood counts, antibodies, hormones, hepatitis, C-reactive protein) describing physiological function. The Medications module houses dietary supplements, non-prescription antacids, prescription medications, and preventive aspirin taken by the participant. The Questionnaire module contains information from self-reported questionnaires on physical activity, general health status, medical conditions, and diabetes. The Occupation module contains information on the participants’ industry and job title. The Weights module contains the survey weights used to make the statistical estimates representative of the US population. The Chemicals module houses information on blood and urine biomarker concentrations of environmental toxicants. The Comments module contains the variables for the chemicals measurements to indicate whether the measurements were above or below the lower limits of detection. While combining the Chemicals and Comments datasets would be natural, we kept these datasets separated to help user differentiate between chemical biomarker variables vs. comments codes to facilitate ease of browsing the data for different needs (e.g., sole interest in the Comments dataset).

To build each dataset, we downloaded 11 Mortality, 36 Dietary, 12 Demographics, 275 Response, 12 Medications, 41 Questionnaire, 10 Occupation, 237 Chemicals, 248 Weights, and 234 Comments data files. Table 1 and Figure 1D show the total number of NHANES files downloaded and the number of variables for each module.

**Table 1.** Number of variables and files for each NHANES type of dataset or module.

| Module | Description | Number of variables | Number of files |
| --- | --- | --- | --- |
| Occupation | Industry and occupational title | 61 | 10 |
| Mortality | Cause of death in 2019 | 11 | 11 |
| Demographics | Demographic attributes | 315 | 12 |
| Medications | Prescription medications | 94 | 12 |
| Dietary | Nutrient information estimated from the food consumption | 649 | 36 |
| Questionnaire | Self-reported questionnaire items (physical activity, general health status, medical conditions, diabetes) | 1267 | 41 |
| Comments | Indicators of the chemical measurements being above or below the lower limit of detection | 469 | 234 |
| Chemicals | Biomarker results of environmental toxicant exposures | 632 | 237 |
| Weights | Survey weights | 252 | 248 |
| Response | Physical examination and physiological measurements | 1045 | 275 |

### 3. Cleaning procedure of each module

We documented inconsistencies in each module for each affected variable (Figure 1E). We tabulated the following common inconsistencies: 1) values indicating “Blank but applicable/cannot be determined”, 2) changes in variable names across cycles for the same variable, 3) changes in measurement units for the same variable, 4) changes in levels for the same category, and 5) multiple replicates instead of one for the same variable. While most laboratory tests are measured once, some laboratory tests such as cotinine were measured at least twice, leading to multiple replicates for the same variable. We also tabulated other inconsistencies that are more specific to the different modules. Once we documented the different types of inconsistencies for each affected variable (Figure 1F), we curated each module by developing a coding pipeline (Figure 1G). In this section, we further detailed the inconsistencies and the curation performed on each module to obtain the harmonized dataset.

The CDC ascertained causes and time to death information by linking eligible participants to the National Death Index. For the Mortality dataset cleaning process, we (1) excluded variables that do not have any data (Table S1) and (2) included additional variables where the categories are described instead of enumerated (Table S2). (1) We excluded variables that have no data recorded as those variables were relevant to the National Health Interview Survey, but not for NHANES. We excluded the quarter of death, year of death, sample weights adjusted for ineligible respondents, and person-level sample weights adjusted for ineligible respondents. In Table S1, we added a note, “excluded due to no data”, in column codename_name to indicate which mortality variables were excluded. (2) We included additional variables to change numeric labels of the categories (e.g., 1 and 0 for mortality status) into readable descriptions (e.g., assumed deceased - 1 and assumed alive - 0). We created labeled variables for eligibility status, mortality status, underlying causes of death, death due to diabetes, and death due to hypertension (Table S2).

The Demographics dataset contains information to describe the characteristics of a representative sample of the US population. (1) We recorded categories indicating “blank but applicable” as missing. “Blank but applicable/cannot be determined” means that the participant was eligible to receive the questionnaire or laboratory test but did not actually receive it due to a variety of reasons: lack of time, low on staff, loss of data, broken container, language barrier, or unreliability. This category is indicated by a series of 8’s, but the number of 8’s differs by variable. For example, “8” indicates “blank but applicable” for “In what country {were you/was SP} born?”, but “88” indicates “blank but applicable” for “Education level - Adults 20+”. In total, we recorded which series of 8’s mean “blank but applicable” for 108 unharmonized demographic variables (column convert_to_NA, Table S3). (2) We checked for the inconsistency of same variable with different names. As an example of the same variable having two names, we use the demographic question: “In what country {were you/was SP} born?”. This response is recorded with four different names in NHANES: HFA6XCR (1988-1994), DMDBORN (1999- 2006), DMDBORN2 (2007-2010), and DMDBORN4 (2011-2018). To ensure that there is only one name for the same variable, we harmonized the name for “In what country {were you/was SP} born?” to be DMDBORN4. In total, we screened for this inconsistency in variable names across 34 demographic factors to produce 26 harmonized variables (columns codename_change and corrected_variable_codename, Table S3). (3) We also checked for the inconsistency: categories for the same variable changing over the study period. This demographic question “In what country {were you/was SP} born?” also enables us to illustrate this problem. Fortunately, one of the categories, “Born in 50 Us states or Washington, DC” (recorded as “1”) is consistent for the entire study period of 1988-2018. Unfortunately, this is not the case for the categories of “Born in Mexico” (1999-2010), “Born in Other Spanish Speaking Country” (2007-2010), “Born in Other Non-Spanish Speaking Country” (2007-2010), “Others” (2011-2018), “Refused” (1999-2018), and “Don’t Know” (1988-2018). “Born in Mexico” was indicated with “2” for 1999-2010, but in the most recent cycles, “2” indicated “Others”. To harmonize the categories across the study period, we collapsed “Born in Mexico”, “Born in Other Spanish Speaking Country”, and “Born in Other Non-Spanish Speaking Country” into one category “Others”, recorded as “2”. The category “Refused” was originally recorded as “7” for 1999-2010 and then was recorded as “77” for 2011-2018, so we harmonized the “Refused” category to be indicated with “77”. Similarly, the category “Don’t Know” was recorded as “9” for 1988-2010 and then was changed to “99” for 2011-2018, so we harmonized the “Don’t Know” to be denoted with “99”. In total, we corrected for the inconsistency in categories for 17 demographic factors (Table S4). Our cleaning process was similar for the Questionnaire dataset (Tables S5 and S6).

The Dietary dataset contains information on estimates of nutrients from food consumed from the 24-hour recall interview as well as dietary behaviors. We checked for (1) the need to record categories indicating “blank but applicable” as missing and (2) the same variable having different names. (3) In addition, we added variables on the summation of fatty acids (Table S7). (4) While the CDC estimated and reported energy, nutrient, and food item intakes on the collection days (e.g., first and second days of intake), these estimates may not be representative of the participants usual dietary consumption habits, so we estimated usual intakes for the NHANES population. (1) “Blank but applicable” are indicated with “8”, “88”, “888”, “8888”, “88888”, “888888”, or “8888888”, which we replaced with NA to denote missingness (column convert_to_NA, Table S7). (2) We corrected for the inconsistency in variable names for 217 dietary factors to produce 143 harmonized variables (column codename_change and corrected_codename, Table S7). (3) We created 7 additional variables for the summation of fatty acids: Sum of SFA 4:0 & 6:0 - Butanoic & Hexanoic (VNDRXS0406), Sum of SFA 8:0, 10:0, & 12:0 - Octanoic, Decanoic, & Dodecanoic (VNDRXS081012), Sum of SFA 14:0 & 16:0 - Tetradecanoic & Hexadecanoic (VNDRXS1416), Sum of SFA 14:0, 16:0, & 18:0 - Tetradecanoic, Hexadecanoic, & Octadecanoic (VNDRXS141618), Total monounsaturated fatty acids (VNDRXSMFAT), Total polyunsaturated fatty acids (VNDRXSPFAT), and Total saturated fatty acids (VNDRXSSFAT). (4) We used the National Cancer Institute method to estimate the usual dietary intakes for each participant. This method is a regression calibration technique, so we inputted the main predictors as the intakes on the first and second days and adjusted for age and sex to predict the usual intakes. Moreover, this procedure requires that the recall interview is repeated. Luckily, the interview was conducted in each NHANES cycle for a total of 11 times. Of note is that the selection of covariates can changed based on the diet-outcome risk model^19^. Therefore, while we provided our usual intakes adjusted for only age and sex, we recommend users to re-estimate the usual intakes based on their disease of interest.

The Medications dataset lists the dietary supplements, non-prescription antacids, prescription medications, and preventive aspirin taken by each participant during a month before the survey. We checked for (1) the need to record categories indicating “blank but applicable” as missing and (2) the same variable having different names. (3) In addition, we included a dictionary to describe the drug codes, which can be found in the file RXQ_DRUG (https://www.n.cdc.gov/Nchs/Nhanes/1999-2000/RXQ_DRUG.htm). (1) “Blank but applicable” are indicated with “8”, “88”, “8888”, “88888”, and “888888”, which we replaced with NA to denote missingness for 8 variables (column convert_to_NA, Table S9). (2) We screened for the inconsistency in variable names across 33 medication variables and corrected 7 affected variables to produce a total of 30 harmonized variables (columns codename_change and corrected_variable_codename, Table S9). (3) In the Medications dataset, multiple medications are recorded with drug codes (RXDDRGID), but such codes are not human readable. Thus, we created an additional column (RXDDRUG) for the drug codes (e.g., “d00116”) to better describe what the drug code means by providing the generic drug name (e.g., “PENICILLIN”). There is a total of 1551 drug codes. We also provide a dictionary (Table S10) that includes more descriptors for the drug codes such as different levels of details on the drug. For example, the first level for the drug category name of “PENICILLIN” is “ANTI-INFECTIVES”, which is the most generic level. The second level is “PENICILLINS”, and the third level is “NATURAL PENICILLINS”. The Medications dataset and the dictionary on the drugs can be merged by the drug codes.

For the Occupation dataset, we checked for (1) inconsistencies in same variable with different names and (2) categories for the same variable changing over time. (3) We included an additional variable to describe the professional job as blue-collar or white-collar and another variable for every combination of industry and collar type. The occupation dataset only contains data from 1999-2014. (1) We screened across 61 occupational variables to ensure that there is only one unique name for the same variable across 33 occupational variables and corrected 5 affected variables to produce a total of 55 harmonized variables (columns codename_change and corrected_variable_codename, Table S11). (2) The categories for the industry and job codes were detailed for 1999-2004 but became more general in the more recent NHANES cycles. For example, the detailed categories include “Waiters and waitresses”, “Cooks”, and “Miscellaneous food preparation and service occupations” for 1999-2004, but these categories were collapsed into “Food Preparation, Serving Occupations” for 2005-2014. For the industry and job codes for the longest and current occupations, we collapse 137 categories into 42 harmonized categories (Table S12). (3) We defined two additional variables by using the job codes from the current job (VNBWCURRJOB) and longest job (VNBWLONGJOB) to categorize the participants as white-collar or blue-collar based on the US Department of Labor definition of blue-collar^22^ (Table S12). We defined another variable for the industry-collar combination (VNSECTORCOLLARCURR). We defined variables that contain the unharmonized categories for the industry and job codes for the longest (VNINDUSTRYLONGOLD – industry and VNLONGJOBOLD – job) and current occupations (VNINDUSTRYOLD – industry and VNCURRJOBOLD – job) (Table S11). These variables can enable researchers to define different methods to collapse the detailed, original industry or job codes into more general categories. We also defined variables that contained the harmonized categories that we used in our previous work on differences in chemical biomarker levels by occupation^8^.

The Chemicals dataset contains biomarker levels of environmental toxicant exposures in the urine or blood. We checked for (1) need to record categories indicating “blank but applicable” as missing, (2) inconsistencies in variable names, (3) units of measurements change over time, and (4) need to harmonize multiple replicates for the same chemical. (5) In addition, we created a few variables to denote the summation of multiple biomarkers. (1) “Blank but applicable” are indicated with “888”, “8888”, “88888”, “888888”, or “8888888”, were replaced with NA to denote missingness (column convert_to_NA, Table S13). (2) There are two examples of inconsistencies in variable names in the Chemicals dataset (columns codename_change and corrected_variable_codename, Table S13): (a) the same variable has many different names or (b) two different variables can have the same name. As an example for case 2a), Triclosan has two variable names, URDTRS and URXTRS. We harmonized the variable name for Triclosan to be URXTRS. As an example for case 2b), 1-(3-Pyridyl)-1-butanol-4-carboxylic acid and 4-hydroxy- 4-(3-pyridyl) butanoic acid are biomarkers for two different chemicals but share the same name, URXHPBT. To differentiate these two chemical biomarkers, we kept the variable name for 1-(3-Pyridyl)-1-butanol-4-carboxylic acid as URXHPBT, and we changed the variable name for 4-hydroxy-4-(3-pyridyl) butanoic acid to VNURXPBA. (3) We corrected the units of measurements for seven chemical biomarkers to ensure that the units are consistent across the entire study period (columns unit_change and converter, Table S13). As an example, chloroform was recorded in pg/mL for 1999-2012, but the units were changed to ng/mL for 2013-2018. We harmonized the units for Chloroform to be pg/mL. Such consistency will also aid in facilitating trend analyses. (4) We use blood cotinine as an example to highlight how we dealt with multiple replicates for the same chemical (columns replicate_codenames and statistic_replicates, Table S13). In 1988-1994, there are three replicates measured for blood cotinine levels, so there are three separate columns in the Chemical datasets to record blood cotinine levels. However, in 1999-2000 and in 2003-2018, LBXCOT is recorded using only one column and is used to denote one replicate measured for blood cotinine levels. To consolidate three columns of cotinine replicates into one column, we calculated the mean of the blood cotinine levels across the three replicates. To harmonize with the measurements of blood cotinine between 1988-1994 and 1999-2018, we recorded this statistic in a column named LBXCOT. Similar to 1988-1994, there are two replicates for blood cotinine levels recorded in 2001-2002 and recorded in two separate columns, one named LBXCOT and another named LB2COT. To consolidate two columns into one column and harmonize the variable name of blood cotinine across the study periods, we calculated the mean of the blood cotinine measured in 2001-2002 and recorded this statistic into a column named LBXCOT. In total, we harmonized 57 replicates into 28 variables. (5) We calculated the summation of Di(2-ethylhexyl) phthalate (DEHP) metabolites by using mono-(2-ethyl-5-oxohexyl) phthalate, mono-(2-ethyl-5- hydroxyhexyl) phthalate, mono-(2-ethylhexyl) phthalate, and mono-2-ethyl-5-carboxypentyl phthalate to by adding the mass weights together and created a variable named VNSUMDEHP. We calculated the summation of arsenic metabolites with monomethylarsonic acid and dimethylarsonic acid and created a variable named VNSUMARS. Measurements for isomers of Perfluorooctane sulfonic acid (PFOS) and Perfluorooctanoic acid (PFOA) were recorded separately in 2013-2018 and for total beta carotene in 2001-2006 and 2017-2018, but the total concentration of these isomers are recorded in the other study periods. To ensure total levels for these mentioned chemical biomarkers are available and consistent across the entire study period, we summed the concentrations of the isomers to calculate the total levels for the mentioned chemical biomarkers, PFOS (LBXPFOS), PFOA (LBXPFOA), and total beta carotene (LBXBCC).

We created the Comments dataset to include the comment codes for the environmental toxicants to indicate whether the chemical concentration is below or above the lower limit of detection (LOD) or far above the range of the assay of the laboratory test. The categories for these comment codes include 0 (above the lower LOD), 1 (below the lower LOD), and 2 (exceeding the calibrated range of the assay). We checked for (1) inconsistencies in variable names, (2) existence of LODs when the chemical was missing a comment code, (3) inconsistency when the comment code exists, but the chemical measurement was missing, and (4) unreasonable comment codes. (1) We ensured that there is only one variable name for each comment codename. We corrected this inconsistency in variable names for 38 chemical comments to produce 25 harmonized chemical comments (column corrected_comment_codename, Table S13). (2) We documented the LODs for as many chemical biomarkers as possible (columns LOD and LOD_notes, Table S13). We created a chemical codename for chemicals with missing comment code but have a recorded LOD. (3) We excluded comment codes when the participant was not measured for the corresponding chemical biomarker. (4) As 37 is not a valid category for the comment code for mono-(2- ethylhexyl) phthalate (URDMHPLC), we corrected the category to indicate whether the measurement of the participants with this mistake are either above or below the LOD. Overall, there is a total of harmonized 502 comment codenames.

The Weights dataset contains the NHANES survey weights to produce estimates that are representative of the US civilian population. We compiled the data files of survey weights and renamed the variables to start with “WT” for consistency in naming between the chemical codenames and their corresponding weight codenames (Table S14). We also created variables that contain the survey weights to be used for each chemical biomarker which enables analysis across several NHANES cycles. For example, to conduct an analysis on blood lead across the entire study period of 1988-2018, we would have to use the following weight variables for a given study period: WTPFEX6 (1988-1994), WTMEC4YR (1999-2002), WTMEC2YR (2003-2012),

WTSH2YR (2013-2016), and WTMEC2YR (2017-2018). To facilitate ease of analysis across multiple study periods, we consolidated all these weight variables for blood lead (“LBXBPB”) in one variable labeled “WT_LBXBPB”. We created a consolidated variable of survey weights for each chemical biomarker. Each consolidated variable is labeled as the concatenation of “WT_” and the variable name of the chemical biomarker measurements.

The Response dataset contains measurements from the physical examination and the laboratory tests that quantify biomarker levels of physiological indicators in urine, blood, or serum. We checked for (1) need to record categories indicating “blank but applicable” as missing, (2) inconsistencies in variable names, (3) categories for the same variable changing over time, and (4) need to harmonize multiple replicates for the same physiological indicator. (5) We also created variables for estimated Glomerular Filtration Rate (eGFR) and the ratios of different types of cholesterol. (1) In the Response dataset, “Blank but applicable” were indicated with “8”, “88”, “888”, “8888”, “88888”, “888888”, or “8888888”, which we replaced with NA to denote missingness. For the variable measuring a type of skinfold, “5555” indicates “skinfold too large for caliper”, which we also replaced with NA to denote missingness (column convert_to_NA, Table S14). (2) An example of inconsistency in variable naming in the Response and Chemicals dataset is two different variables having the same variable name. For example, serum Hydroxycotinine (ng/mL) and Hematocrit (%) are different biomarkers but have the same variable name, LBXHCT. To differentiate the two biomarkers, we change the variable name for Hematocrit (%) to VNHEMACRIT (columns codename_change and corrected_variable_codename, Table S14). (3) The categories to indicate why waist circumference could not be measured were detailed in 1988-1994 (e.g., “Refused” and “Crying/fighting/upset/uncooperative”) and then became more general in 1999-2018 (e.g., “Could not obtain”). We harmonized all categories to be “Could not obtain” (Table S16). (4) Our procedure to collapse multiple replicates into one variable for the Response dataset is the same as in the Chemicals dataset. In total, we harmonized 429 replicates into 206 variables. (5) We created two estimates of GFR with one adjusting for race (VNEGFRADJ) and the other not adjusting for race (VNEGFR). We use the most widely used equation for GFR, Chronic Kidney Disease Epidemiology Collaboration (CKD-EPI) equation^23^. In addition, we created two variables for cholesterol, one for the ratio of LDL to HDL Cholesterol (VNLDHDLRATIO) and another for the ratio of Total to HDL Cholesterol (VNTOTHDRATIO) (Table S14).

Finally, we combined the 614 separate files to create the following individual, curated datasets: 1) Mortality, 2) Demographics, 3) Questionnaire, 4) Dietary, 5) Medications, 6) Occupation, 7) Chemicals, 8) Comments, 9) Weights, and 10) Response. We merged by “SEQN”, “SEQN_new”, and “SDDSRVYR” to create these datasets. There is a grand total of 4,740 variables and 134,310 participants. Figure 1H shows the number of variables and participants by cleaned module.

### 4. Creating the data dictionary

We created a data dictionary that lists the variable names and other descriptors to help facilitate analysis and scanning through the datasets (Figure 1I and 1J). In this data dictionary, we list the variable name (e.g., BMXBMI), readable description (e.g., Body Mass Index (kg/m^2^)), module (e.g., Response), category (e.g., Body Measures), total number of participants with measurements (e.g., 115,629 participants with measurements for BMI), and survey cycles with measurements (e.g., BMI was measured in 1988-1994 and 1999-2018). Second, we further classified the variables into more specific subcategories. These subcategories provide higher resolution compared to the module to further help browse the data. For example, the more specific category for BMI is “Body Measures”. Table 2 displays the number of variables for each combination of module and category. Third, for variables in the Chemicals dataset, we included measurements units, CAS Number, comment codename (variable from the Comments dataset), name of the chemical family, and abbreviated version of the chemical family. CAS Number, also known as CAS registry number, is a unique numeric identifier assigned to every chemical substance found in the open scientific literature^24^. The chemical biomarkers in NHANES are not recorded with any CAS numbers, so we provided the CAS Numbers for 429 toxicants to help facilitate linkage between the NHANES chemical exposures data with other toxicological databases. Fourth, we linked the comment codenames to their corresponding chemical biomarker to help users identify which chemical concentrations are above or below the lower LOD. Finally, we categorized the chemical biomarkers into their chemical classes based on chemical functions or structural properties. The entire dictionary is provided as Table 3.

**Table 2.** List of specific categories, corresponding module or NHANES type of dataset, and an number of variables in each category.

| Category | Module | Number of variables |
| --- | --- | --- |
| Acrylamide & Glycidamide | Chemicals | 2 |
| Aldehydes | Chemicals | 12 |
| Aromatic Amines | Chemicals | 6 |
| Aromatic Diamines | Chemicals | 5 |
| Atrazine and Metabolites | Chemicals | 6 |
| Brominated Flame Retardants | Chemicals | 22 |
| Cadmium | Chemicals | 4 |
| Calcium | Chemicals | 3 |
| Chloride | Chemicals | 3 |
| Chromium & Cobalt | Chemicals | 2 |
| Copper | Chemicals | 1 |
| Cotinine | Chemicals | 16 |
| DEET and Metabolites | Chemicals | 4 |
| Dioxins, Furans, & Coplanar PCBs | Chemicals | 116 |
| Environmental Pesticides | Chemicals | 5 |
| Ethylene Oxide | Chemicals | 1 |
| Flame Retardant Metabolites | Chemicals | 12 |
| Formaldehyde | Chemicals | 1 |
| Heterocyclic Aromatic Amines | Chemicals | 10 |
| Inorganic Mercury | Chemicals | 3 |
| Iodine | Chemicals | 3 |
| Lead | Chemicals | 3 |
| Manganese | Chemicals | 1 |
| Melamine | Chemicals | 2 |
| Mercury | Chemicals | 2 |
| Methylmalonic Acid | Chemicals | 1 |
| Neonicotinoids | Chemicals | 6 |
| Nitrosamines | Chemicals | 3 |
| Non-persistent Pesticide Metabolites | Chemicals | 25 |
| Organophosphate Insecticides - Diakyl Phosphate Metabolites | Chemicals | 6 |
| Perchlorate, Nitrate & Thiocyanate | Chemicals | 3 |
| Perfluoroalkyl and Polyfluoroalkyl Substances | Chemicals | 31 |
| Personal Care and Consumer Product Chemicals and Metabolites | Chemicals | 11 |
| Pesticides - Carbamates & Organophosphorus Metabolites | Chemicals | 6 |
| Pesticides - Organochlorine Metabolites | Chemicals | 25 |
| Phthalates & Plasticizers Metabolites | Chemicals | 21 |
| Phytoestrogens | Chemicals | 6 |
| Polycyclic Aromatic Hydrocarbons | Chemicals | 12 |
| Potassium | Chemicals | 3 |
| Pyrethroids, Herbicides, & Organophosphorus Metabolites | Chemicals | 11 |
| Selenium | Chemicals | 3 |
| Sodium | Chemicals | 3 |
| Terephthalate | Chemicals | 2 |
| Total & Speciated Arsenics | Chemicals | 9 |
| Urinary Metals | Chemicals | 15 |
| Vitamin A, Vitamin E & Carotenoids | Chemicals | 44 |
| Vitamin B | Chemicals | 5 |
| Vitamin C | Chemicals | 3 |
| Vitamin D | Chemicals | 6 |
| Volatile N-Nitrosamine Compounds | Chemicals | 5 |
| Volatile Organic Compound (VOC) Metabolites | Chemicals | 84 |
| Zinc | Chemicals | 1 |
| Acrylamide & Glycidamide | Comments | 2 |
| Aldehydes | Comments | 12 |
| Aromatic Amines | Comments | 6 |
| Aromatic Diamines | Comments | 5 |
| Atrazine and Metabolites | Comments | 6 |
| Brominated Flame Retardants | Comments | 11 |
| Cadmium | Comments | 4 |
| Calcium | Comments | 3 |
| Chloride | Comments | 3 |
| Chromium & Cobalt | Comments | 2 |
| Copper | Comments | 1 |
| Cotinine | Comments | 16 |
| DEET and Metabolites | Comments | 4 |
| Dioxins, Furans, & Coplanar PCBs | Comments | 58 |
| Environmental Pesticides | Comments | 5 |
| Ethylene Oxide | Comments | 1 |
| Flame Retardant Metabolites | Comments | 12 |
| Formaldehyde | Comments | 1 |
| Heterocyclic Aromatic Amines | Comments | 10 |
| Inorganic Mercury | Comments | 3 |
| Iodine | Comments | 3 |
| Lead | Comments | 3 |
| Manganese | Comments | 1 |
| Melamine | Comments | 2 |
| Mercury | Comments | 2 |
| Methylmalonic Acid | Comments | 1 |
| Neonicotinoids | Comments | 6 |
| Nitrosamines | Comments | 3 |
| Non-persistent Pesticide Metabolites | Comments | 25 |
| Organophosphate Insecticides - Diakyl Phosphate Metabolites | Comments | 6 |
| Perchlorate, Nitrate & Thiocyanate | Comments | 3 |
| Perfluoroalkyl and Polyfluoroalkyl Substances | Comments | 31 |
| Personal Care and Consumer Product Chemicals and Metabolites | Comments | 11 |
| Pesticides - Carbamates & Organophosphorus Metabolites | Comments | 6 |
| Pesticides - Organochlorine Metabolites | Comments | 13 |
| Phthalates & Plasticizers Metabolites | Comments | 21 |
| Phytoestrogens | Comments | 6 |
| Polycyclic Aromatic Hydrocarbons | Comments | 12 |
| Potassium | Comments | 3 |
| Pyrethroids, Herbicides, & Organophosphorus Metabolites | Comments | 11 |
| Selenium | Comments | 3 |
| Sodium | Comments | 3 |
| Terephthalate | Comments | 2 |
| Total & Speciated Arsenics | Comments | 9 |
| Urinary Metals | Comments | 15 |
| Vitamin A, Vitamin E & Carotenoids | Comments | 32 |
| Vitamin B | Comments | 5 |
| Vitamin C | Comments | 3 |
| Vitamin D | Comments | 6 |
| Volatile N-Nitrosamine Compounds | Comments | 5 |
| Volatile Organic Compound (VOC) Metabolites | Comments | 84 |
| Zinc | Comments | 1 |
| Demographics | Demographics | 281 |
| Breast-feeding | Dietary | 1 |
| Diet Behaviors | Dietary | 25 |
| Dietary Interview | Dietary | 8 |
| Fish & Shellfish | Dietary | 66 |
| Food Patterns Equivalents | Dietary | 37 |
| Salt | Dietary | 6 |
| Survey Variables | Dietary | 6 |
| Total Nutrient Intakes | Dietary | 164 |
| Water | Dietary | 5 |
| Mortality | Mortality | 16 |
| Occupation | Occupation | 70 |
| Alcohol Use | Questionnaire | 15 |
| Current Health Status | Questionnaire | 13 |
| Diabetes | Questionnaire | 89 |
| Diet | Questionnaire | 38 |
| Exercise | Questionnaire | 35 |
| Family Background | Questionnaire | 17 |
| Food Frequency | Questionnaire | 81 |
| Health Insurance and Income Assessment | Questionnaire | 36 |
| Medical Conditions | Questionnaire | 579 |
| Physical Activity | Questionnaire | 113 |
| Prescription Medications | Questionnaire | 87 |
| Tobacco | Questionnaire | 30 |
| Vitamin, Mineral, and Medicine Usage | Questionnaire | 192 |
| Alanine aminotransferase | Response | 4 |
| Albumin | Response | 10 |
| Alkaline phosphatase | Response | 3 |
| Antibody | Response | 17 |
| Apolipoprotein | Response | 12 |
| Aspartate aminotransferase | Response | 3 |
| Bicarbonate | Response | 3 |
| Bilirubin | Response | 7 |
| Bioelectrical Impedance Analysis | Response | 124 |
| Blood Pressure | Response | 75 |
| Blood Urea Nitrogen | Response | 6 |
| Body Measures | Response | 127 |
| Bone Alkaline Phosphatase | Response | 3 |
| C-reactive protein | Response | 3 |
| Cardiovascular Fitness | Response | 65 |
| CD8+ T Cells | Response | 2 |
| Cholesterol | Response | 40 |
| Complete Blood Count | Response | 150 |
| Creatine Phosphokinase | Response | 1 |
| Creatinine | Response | 6 |
| Cytomegalovirus | Response | 2 |
| Cytomegalovirus IgG & IgM Antibodies | Response | 6 |
| Diabetes | Response | 6 |
| Epstein-Barr Virus | Response | 2 |
| Erythrocyte Protoporphyrin | Response | 6 |
| Ferritin | Response | 6 |
| Fibrinogen | Response | 6 |
| Folate | Response | 12 |
| Follicle Stimulating Hormone | Response | 7 |
| Gamma Glutamyl Transferase | Response | 4 |
| Globulin | Response | 6 |
| Glomerular Filtration Rate | Response | 2 |
| Glycohemoglobin | Response | 3 |
| Hepatitis | Response | 18 |
| Herpes Simplex Virus Type-1 & Type-2 | Response | 6 |
| High-Sensitivity C-Reactive Protein | Response | 3 |
| HIV | Response | 4 |
| Homocysteine | Response | 3 |
| Iron | Response | 21 |
| Lactate dehydrogenase | Response | 3 |
| Lipoprotein | Response | 6 |
| Luteinizing Hormone | Response | 6 |
| Measles, Rubella, & Varicella | Response | 11 |
| Normalized Calcium | Response | 3 |
| Oral Glucose Tolerance Test | Response | 8 |
| Osmolality | Response | 3 |
| Phlebotomy Screening | Response | 21 |
| Phosphorus | Response | 6 |
| Plasma Fasting Glucose & Insulin | Response | 30 |
| Serum C-peptide | Response | 10 |
| Sex Steroid Hormone | Response | 1 |
| Spirometry | Response | 103 |
| Thyroid Profile | Response | 13 |
| Total Protein | Response | 6 |
| Transferrin | Response | 1 |
| Uric Acid | Response | 6 |
| Urinary N-telopeptides | Response | 3 |
| Survey Weights | Weights | 262 |
| Survey Weights - Chemicals | Weights | 595 |

For categorical variables, we created Table 4 to display the levels or categories and their corresponding readable descriptions. We only list the categorical variables for which the categories were harmonized.

### 5. Starter code for mortality-related exposome analysis

We created four R markdown files to help users conduct exposome analysis: “example_0 - merge_datasets_together.Rmd”, “example_1 - account_for_nhanes_design.Rmd”, “example_2 - calculate_summary_statistics.Rmd”, and “example_3 - run_multiple_regressions.Rmd”. We recommend going through the tutorials in order. The tutorial on merging datasets demonstrates how to merge the curated NHANES datasets together. The tutorial on accounting for NHANES sampling design goes through how to conduct a linear regression model, a survey-weighted regression model, a Cox proportional hazard model, and a survey-weighted Cox proportional hazard model. The tutorial on summary statistics will guide the user on how to calculate summary statistics for one variable with and without accounting for the sampling design. Then the tutorial will guide the users to expand such statistics to multiple variables. The tutorial on running multiple regression models guides the user through how to first run conduct a linear regression model and a survey-weighted regression model for lead, and then expand the methodology to 37 metal biomarkers. This tutorial also focuses on how to use the survey weights that we created for each chemical biomarker.

## Data Records

### Data record 1

Curated NHANES datasets and data dictionary in .csv format and .RData format along with starter code for exposome analysis The curated NHANES datasets and the data dictionaries includes 13 .csv files and 1 excel file. The curated NHANES datasets involves 10 .csv formatted files, one for each module and labeled as the following: 1) mortality, 2) dietary, 3) demographics, 4) response, 5) medications, 6) questionnaire, 7) chemicals, 8) occupation, 9) weights, and 10) comments. The 11^th^ file is a dictionary that lists the variable name, description, module, category, units, CAS Number, comment use, chemical family, chemical family shortened, number of measurements, and cycles available for all 4,740 variables in NHANES (“dictionary_nhanes.csv”). The 12^th^ csv file contains the harmonized categories for the categorical variables (“dictionary_harmonized_categories.csv”). The 13^th^ file contains the dictionary for descriptors on the drugs codes (“dictionary_drug_codes.csv”). The 14^th^ file is an excel file that contains the cleaning documentation, which records all the inconsistencies for all affected variables to help curate each of the NHANES datasets (“nhanes_inconsistencies_documentation.xlsx”).

For researchers who want to conduct their analysis in the R programming language, the curated NHANES datasets and the data dictionaries can be downloaded as a .zip file which include an .RData file and an .R file. We provided an .RData file that contains all the aforementioned datasets as R data objects (“w - nhanes_1988_2018.RData”). Also in this .RData file, we make available all R scripts on customized functions that were written to curate the data. We also provide an .R file that shows how we used the customized functions (i.e. our pipeline) to curate the data (“m - nhanes_1988_2018.R”).

The starter codes for exposome analysis are available as an R markdown file (.Rmd): “example_0 - merge_datasets_together.Rmd”, “example_1 - account_for_nhanes_design.Rmd”, “example_2 – calculate_summary_statistics.Rmd”, and “example_3 - run_multiple_regressions.Rmd”.

### Data record 2

Curated NHANES datasets and data dictionary on figshare All formats listed in Data record 1 are also available on figshare (https://figshare.com/articles/dataset/NHANES_1988-2018/21743372). Figshare is a platform publishing data that is free for all researchers.

### Data record 3

Curated NHANES datasets and data dictionary on Kaggle All formats listed in Data record 1 are also available on Kaggle (https://www.kaggle.com/datasets/nguyenvy/nhanes-19882018?select=dictionary_nhanes.csv). Kaggle is an online community platform for data scientists and machine learning enthusiasts. The platform provides opportunities for collaboration among users, browsing and publication of datasets, access to GPU integrated notebooks, and competition against other scientists to solve data science challenges.

### Data record 4

Curated NHANES datasets and data dictionary on Hugging Face All formats listed in Data record 1 are also available on Hugging Face

(https://huggingface.co/datasets/nguyenvy/cleaned_nhanes_1988_2018). Hugging Face is an AI community with goals to build, train, deploy, and democratize machine learning models and freely available datasets.

### Data record 5

Curated NHANES datasets and data dictionary on Google Dataset

All formats listed in Data record 1 are also available on Google Dataset Search (https://datasetsearch.research.google.com/search?src=0&query=NHANES%201988- 2018&docid=L2cvMTF0bm1zY2xqeA%3D%3D). This record is a link to the data record on Kaggle. Google Dataset Search is a search engine from Google that help researchers find freely available online data.

### Technical Validation

We obtained the raw data from the CDC NHANES. Quality control and quality assurance protocols to describe the validation of their data are provided in laboratory procedure manuals.

## Usage Notes

### Code to merge the curated NHANES datasets

The curated NHANES datasets can be merged by “SEQN”, “SEQN_new”, and “SDDSRVYR”. We provide code in our GitHub repository and all data records to merge the datasets together (“example_0 - merge_datasets_together.Rmd”).

### NHANES sampling design

The CDC deployed a multistage sampling design to oversample minority groups such as adolescents, older Americans, Mexican Americans, Non-Hispanic Black Americans, pregnant women, and other Americans at or below 130% of the poverty income line. We caution that statistical analyses need to account for the NHANES sampling design to enable estimates such as means and standard errors, to be representative of the non-institutionalized, civilian US population. We provide a R file in our GitHub repository and all data records to demonstrate how to account for the NHANES sampling design when analyzing survey data via running a generalized linear model and a Cox proportional hazards model (“example_1 - account_for_nhanes_design.Rmd”) along with calculating summary statistics (“example_2 - calculate_summary_statistics.Rmd”).

Exposome-wide association approaches for analyzing mortality and secular trends tidyverse is a collection of packages in R designed for data science. We have used functions from this package to obtain summary statistics over multiple variables and run multiple regression models with writing the fewest lines of code. We provide code in our GitHub repository and all data records to help facilitate exposome-wide approaches to analyzing the NHANES data (“example_3 - run_multiple_regressions.Rmd”).

### Long versus wide format of NHANES datasets

We provided the individual datasets in different forms: long vs. wide. The dietary and medications datasets are long (i.e., there are multiple rows for the same participant), while the other datasets are wide (i.e., there is only one row per participant). We caution merging long datasets with wide dataset as this will lead to duplicates of the participant information, and in turn, huge consumption of data storage.

### Uncleaned and cleaned NHANES datasets

There are two set of datasets for each of the modules: the uncleaned, raw dataset and the curated dataset. Please be careful when use our curation strategy as there may be inconsistencies or mistakes that we are not aware of.

### Replicates of the measurements

We calculated the mean concentration across the replicates for the same chemical biomarker to consolidate the multiple replicates into one. We provide the replicates separately if researchers prefer to calculate other statistics instead of the mean.

## Data Availability

All data produced are available online at figshare, Kaggle, Hugging Face, and Google Dataset.

https://figshare.com/articles/dataset/NHANES_1988-2018/21743372

https://www.kaggle.com/datasets/nguyenvy/nhanes-19882018?select=dictionary_nhanes.csv

https://huggingface.co/datasets/nguyenvy/cleaned_nhanes_1988_2018

https://datasetsearch.research.google.com/search?src=0&query=NHANES%201988-2018&docid=L2cvMTF0bm1zY2xqeA%3D%3D

## Code Availability

Our curation code is publicly available on GitHub (https://github.com/vynguyen92/harmonized_nhanes_1988_2018.git).

## Author Contributions

Vy Kim Nguyen: Conceptualization, Software, Data curation, Visualization, Writing – original draft, Writing – review & editing, Project administration, Supervision

Lauren Y. M. Middleton: Conceptualization, Data curation, Writing – review & editing Lei Huang: Writing – review & editing, Data curation

Neil Zhao: Data curation, Writing – review & editing Eliseu Verly Jr.: Data curation, Writing – review & editing Jacob Kvasnicka: Software, Data curation

Luke Sagers: Conceptualization, Writing – review & editing

Chirag J. Patel: Conceptualization, Writing – review & editing, Resources, Supervision Justin Colacino: Conceptualization, Writing – review & editing, Resources

Olivier Jolliet: Conceptualization, Writing – review & editing, Funding acquisition, Supervision, Project administration, Resources

## Competing Interest

All authors report no conflicts of interest.

Table 3. Data dictionary listing the variable name, description, module, category, units, CAS Numbers, comment, chemical family, number of measurements, and cycles with measurements. CAS Numbers, comment use, and chemical family are descriptors for variables on environmental exposure toxicants. Comment use indicates which comment code to use to identify which chemical concentration are above or below the lower LOD.

[This table is in an excel file because there are >5000 rows.]

Table 4. Harmonized levels for categorical variables. [This table is in an excel file because there are 486 rows.]

